# Aripiprazole as a candidate treatment of COVID-19 identified through genomic analysis

**DOI:** 10.1101/2020.12.05.20244590

**Authors:** Benedicto Crespo-Facorro, Miguel Ruiz-Veguilla, Javier Vázquez-Bourgon, Ana C. Sánchez-Hidalgo, Nathalia Garrido-Torres, Jose M. Cisneros, Carlos Prieto, Jesus Sainz

## Abstract

**Background:** Antipsychotics suppress expression of inflammatory cytokines and inducible inflammatory enzymes. Elopiprazole (a phenylpiperazine antipsychotic drug in phase 1) has been characterized as a therapeutic drug to treat SARS-CoV-2 infection in a repurposing study. We aim to investigate the potential effects of aripiprazole (an FDA approved phenylpiperazine) on COVID19-related immunological parameters.

**Methods:** Differential gene expression profiles of non-COVID versus COVID RNA-Seq samples (CRA002390 project in GSA database) and drug-naïve patients with psychosis at baseline and after three months of aripiprazole treatment was identified. An integrative analysis between COVID and aripiprazole immunomodulatory antagonist effects was performed.

**Findings:** 82 out the 377 genes (21.7%) with expression significantly altered by aripiprazole have also their expression altered in COVID-19 patients and in 93.9% of these genes their expression is reverted by aripiprazole. The number of common genes with expression altered in both analyses is significantly higher than expected (Fisher’s Exact Test, two tail; P value=3.2e-11). 11 KEGG pathways were significantly enriched with genes with altered expression both in COVID-19 patients and aripiprazole medicated schizophrenia patients (P adj<0.05). The most significant pathways were associated to the immune system such as the “inflammatory bowel disease (IBD)” (the most significant pathway with a P adj of 0.00021), “Th1 and Th2 cell differentiation” and “B cell receptor signaling pathway”, all three related to the defense against infections.

**Interpretation:** This exploratory investigation may provide further support to the notion that protective effect is exerted by phenylpiperazine by modulating the immunological dysregulation associated to COVID-19. Along with many ongoing studies and clinical trials, repurposing available medications could be of use in countering SARS-CoV-2 infection, but require further studies and trials.

## 1. Introduction

The SARS-CoV-2 epidemic has become the greatest challenge facing medicine today. Infected patients present with a wide range of clinical severity varying from asymptomatic to fatal condition (Wu et al., 2020). Advanced age, gender (male) and suffering comorbidities (diabetes, cardiovascular or chronic respiratory diseases) are risk factors for higher clinical severity, hospitalization rate and death from COVID-19 (Rubino et al., 2020; Zhou et al., 2020).

The presence of these comorbidities may decrease resilience and lower the ability to tolerate additional cytokine storm (Mangalmurti & Hunter, 2020). The high prevalence of these comorbidities among individuals with severe mental illnesses should increase the risk of COVID-19 complications and therefore might prone to a poorer outcome. However, somewhat unexpectedly low incidence and lack of severity of SARS-CoV-2 infection among people with schizophrenia have been reported (de Lusignan et al., 2020; van der Meer et al., 2020).

COVID-19 individuals who become critically and fatally ill seem to experience an indiscriminate and runaway immune response with an unchecked systemic overproduction of cytokines and immunological disbalance(Bhaskar et al., 2020; Manjili et al., 2020; Zeng et al., 2020).

Although, COVID-19 related immunopathogenesis is not understood just as an emergent cytokine storm but also as an impairment of protective T cell immunity (Chen et al., 2020).

Anti-inflammatory drugs such as dexamethasone have been shown to reduce deaths (Vabret et al., 2020). The crucial role of NLRP3 inflammasome activation in the pathogenesis of diseases caused by SARS-CoVs draws also attention toward potential role of its inhibitors in the treatment of COVID-19 (van den Berg & Te Velde, 2020).

The last decade has drastically increased the evidence linking psychosis spectrum disorders and altered immune function (Radhakrishnan et al., 2017). An imbalance in the host immune response seems to be associated with the activation of microglia in the pathophysiology of schizophrenia (Barichello et al., 2020). Antipsychotics were largely reported to suppress expression of inflammatory cytokines and inducible inflammatory enzymes (i.e., cyclooxygenase) and microglia activation (Dinesh et al., 2020). These anti-inflammatory effects are elicited via the reduction of proinflammatory cytokines production, modulating monocytes response through TLR and the inhibition of the microglial activation by reducing the levels of inducible nitric oxide synthase (iNOS), IL-1β, IL-6, and TNF-α^16-18^. The immunomodulatory effect of risperidone (pyridopyrimidines) and aripiprazole (marketed phenylpiperazine) has been demonstrated (Juncal-Ruiz et al., 2018), although aripiprazole had greater anti-inflammatory effect on TNF-α, IL-13, IL-17α and fractalkine. Wide range of different drug classes, such as cancer therapeutics, antipsychotics, and antimalarials, seem to have a beneficial effect against MERS and SARS coronaviruses (Dyall et al., 2017). A recent paper (Weston et al., 2020) described that, although infection cannot be prevented, chlorpromazine protects mice from severe clinical disease and SARS-CoV. Clozapine (atypical antipsychotic) has revealed to be effective in suppressing the proinflammatory cytokine expression by limiting the NLRP3 inflammasome activation in an in vitro model of schizophrenia (Giridharan et al., 2020). Interestingly, a research(L. Riva et al., 2020) investigating approximately 12,000 drugs in clinical-stage or Food and Drug Administration (FDA)-approved small molecules to identify candidate drugs to treat COVID-19, reported that elopiprazole (a never marketed phenylpiperazine antipsychotic drug) was listed among the 21 most potent compounds to inhibit SARS-CoV infection. Thus, the protective effect of phenylpiperazine marketed antipsychotics (aripiprazole) against a pernicious cytokine storm is a hypothesis that warrants further investigation.

The aim of the present study was to examine the potential immunomodulatory effects of antipsychotics by analyzing the profile of gene expression of drug-naïve patients with psychosis at baseline and after three months of treatment with aripiprazole (a phenylpiperazine antipsychotic drug).

## 2 Materials and Methods

### 2.1 Setting and Sample study

The cohort analyzed to study the effect of aripiprazole was obtained at the University Hospital Marques de Valdecilla (Cantabria, Spain). Conforming to international standards for research ethics, this study was approved by the Cantabria Ethics Institutional Review Board (IRB). Patients meeting inclusion criteria for a first episode of psychosis (drug-naïve) and their families provided written informed consent to be included in the study. After informed consent was signed, patients were included in a prospective, randomized, flexible-dose, open-label study (Crespo-Facorro et al., 2017; Mayoral Van-Son et al, 2020).

### 2.2 Laboratory assessments

Blood samples were obtained from 57 fasting schizophrenia subjects from 8:00 to 10:00 a.m. by the same staff and in the same setting. A detailed description of methodology followed to assess biochemical variables is available under request. None of the patients had a chronic inflammation or infection, or were taking medication that could influence the results of blood tests.

### 2.3 RNA extraction

Total RNA was extracted from blood using the Tempus™ Blood RNA Tube and the Tempus™ Spin RNA Isolation Kit (Applied Biosystems, Foster City, CA, USA) following the manufacturer’s protocols. To select only high-quality RNA, the RNA integrity number (RIN) was characterized with a Bioanalyzer (Agilent Technologies, Santa Clara, CA, USA) and samples with a RIN of at least 7.6 were used.

### 2.4 RNA next-generation sequencing

Total RNA was extracted from peripheral blood of each individual. The messenger RNA (mRNA) obtained from blood was sequenced at the Centro Nacional de Análisis Genómico (CNAG) using Illumina HiSeq instruments (San Diego, CA, USA). The mRNA was isolated from the total RNA and was fragmented once transformed into complementary DNA (cDNA). Fragments of 300bp on average were selected to construct the cDNA libraries for sequencing. Pair-end sequences of 70 nucleotides for each end were produced. The mRNA from blood samples of 57 drug-naïve psychotic patients at baseline and after 3 months of continuous treatment with aripiprazole was sequenced.

Sequence files were aligned to the GRCh38 human reference genome (Gencode release 25) using the STAR aligner(Dobin et al., 2013; Harrow et al., 2012). Reads count were normalized with the Voom algorithm using the cyclic loess method (Law et al., 2014) and significant gene expression changes between treated and naïve patients were identified with limma(Smyth et al., 2005). We performed a paired analysis using a Wald test and a parametric fit type with an adjusted p-value cutoff of 0.01.

### 2.5 Wuhan COVID19 dataset

COVID data was downloaded from the GSA server(Wang et al., 2017) with the CRA002390 identifier (Xiong et al., 2020). This research collected RNA-Seq samples of peripheral blood mononuclear cells (PBMC) from three COVID-19 patients and three healthy donors that were included in this study. These data were analyzed with RaNA-Seq (Prieto & Barrios, 2019) cloud platform and differential expression genes were detected by means of DESeq2 (Love et al., 2014) cloud using a Wald test, a parametric fit type and setting an adjusted p-value cutoff of 0.01.

### 2.6 Statistical and bioinformatic Analysis

Statistical significance of differential expression co-occurrence between COVID and Aripiprazole studies was calculated with a Fisher Exact test. Functional enrichment analyses, to identify biological pathways in the Biosystem database (downloaded on January 2020)(Geer et al., 2010) with a significant presence of differential expressed genes, were carried out with a Fisher Exact test. Visualization and final representation of pathways were performed with the pathview R package(Luo & Brouwer, 2013).

## 3 Results

Integrative transcriptomic analyses of aripiprazole effects on differentially expressed genes in COVID-19

### 3.1 COVID-19 patients (Wuhan dataset) versus controls

We found 2,137 genes with significant differential expression between COVID-19 patients and controls (P adj value <0.05). The most significant gene was the *Charcot-Leyden crystal galectin* gene (*CLC*) with a P adj value of 7.8e-23.

### 3.2 Drug-naïve schizophrenia patients at entry and after 3 months of aripiprazole treatment

We found 377 genes with significant differential expression before and after medication (P adj value <0.05). The two most significant genes were the *LIM domain only 4* (*LMO4*) and the *ATP binding cassette subfamily A member 9* (*ABCA9*) with a P adj value of 0.0039.

### 3.3 KEGG pathways significantly enriched with common genes with altered expression in COVID-19 patients and schizophrenia patients medicated with aripiprazole

We found that 11 KEGG pathways were significantly enriched with genes with altered expression both in COVID-19 patients and aripiprazole medicated schizophrenia patients (P adj<0.05). The most significant pathway was the Inflammatory bowel disease (IBD) with a P adj of 0.00021.

## 4 Discussion

Counterintuitively, the prevalence of psychosis among severely ill SARS-CoV-2 infected individuals is reduced. Severe COVID-19 infection requiring hospitalization does not appear to be associated to schizophrenia spectrum disorders (de Lusignan et al., 2020; van der Meer et al., 2020). Diverse speculative explanations such as higher degree of social isolation among psychosis individuals have been proposed to justify this lack of cases of COVID-19 among this population. The fact that antipsychotics have a demonstrated effect on immunological pathway may lead to the speculation about the beneficial effects of this drugs on controlling the acute hyper-inflammatory response that may be responsible for critical illness(Dinesh et al., 2020).

The present comparison of the genes with altered expression in COVID-19 patients versus controls and schizophrenia patients before and after medication with aripiprazole reveals that 82 out the 377 genes (21.7%) altered by aripiprazole medication are significantly altered also in the cohort of COVID-19 patients. The number of common genes to both analyses is significantly higher than expected by chance (Fisher’s Exact Test, two tail; P value=3.2e-11).

Interestingly, out of the 82 genes with expression altered in both analyzed cohorts 55 genes have decreased expression after aripiprazole medication and increased expression in COVID-19 patients; also, out of the 82 genes common to both cohorts 22 have increased expression after aripiprazole medication and decreased expression in COVID-19 patients. In total 77 genes out of 82 (93.9%) have altered expression in different direction when we compared the effects of COVID-19 and aripiprazole medication, suggesting that aripiprazole may have an effect that reverse many of the changes in expression caused by the coronavirus infection.

Analysis of pathways for enrichment of the 77 common genes with altered expression in COVID-19 and aripiprazole medicated patients shows 11 pathways significantly enriched (P value Fisher <0.05). Several of those pathways are related to the immune system such as the “inflammatory bowel disease (IBD)” (the most significant pathway), “Th1 and Th2 cell differentiation” and “B cell receptor signaling pathway”, both related to the defense against infections. Based on actual evidence showing hyperinflammation as well as T cell deficiencies and coagulation abnormalities, associated with life-threatening organ dysfunction, rather than with a mere hyper-inflammatory disease (Elhusseiny et al., 2020; G. Riva et al., 2020).

COVID-19 individuals who become critically and fatally ill experience an unchecked systemic overproduction of cytokines (Cytokines storm) and immuno-pathology has been suggested as a primary driver of morbidity and mortality with COVID-19 (Vabret et al., 2020). In addition to anti-inflammatory drugs (e.g., dexamethasone) other strategies including repurposing available medications aimed at quelling the inflammatory storm in critically ill patients (Guy et al., 2020).

A recent study (L. Riva et al., 2020) profiled a library of approximately 12,000 drugs in clinical-stage or FDA-approved small molecules to identify candidate drugs to treat COVID-19. In the list of the 21 most potent compounds to inhibit infection validated in dose response across multiple cell lines there are two antipsychotic drugs, elopiprazole and 8-(3-Chlorostyryl) caffeine which are in phase I and preclinical stages respectively. Elopiprazole and aripiprazole belong to the class of organic compounds known as phenylpiperazines. Phenylpiperazines are compounds containing a phenylpiperazine skeleton, which consists of a piperazine bound to a phenyl group.

## Conclusions

The global effort to find therapies and develop vaccines capable of stopping the spread, and end the COVID-19 pandemic is a priority. This combined observational and exploratory investigations may provide further support to the notion that protective effect exerted by phenylpiperazines by modulating the immunological dysregulation associated to COVID-19. Along with many ongoing studies and clinical trials, repurposing available medications could be of use in countering SARS-CoV-2 infection, but require further studies and trials.

## Data Availability

The cohort to study the effect of aripiprazole on gene expression was obtained from Cantabria Biobank. The patterns of gene expression of non-COVID and COVID were obtained from CRA002390 project in GSA database

## Acknowledgements

We are highly indebted to the participants and their families for their cooperation in this study. We also thank IDIVAL biobank (Inés Santiuste and Jana Arozamena) for clinical samples and data as well as the PAFIP members (Marga Corredera) for the data collection. This work was supported by: SAF2016-76046-R and SAF2013-46292-R (MINECO and FEDER) to B.C.F.

We kindly thank all clinical staff at the Hospital Universitario Virgen del Rocio for support to collect clinical records and provide clinical care to COVID-19 patients.

We also kindly thank Dra. Marisa Barrigon for helpful discussions regarding clinical data analysis, and Idalino Rocha for manuscript editing and formatting.

## Author’s contributions

Study concept and design: BC-F, MR-V, CP and JS; acquisition, analysis or interpretation of data: all authors; drafting of the manuscript: all authors; critical revision of the manuscript for important intellectual content: all authors; obtained funding: BC-F and JS; study supervision: BC-F, MR-V, CP and JS.

## References

Barichello, T., Simoes, L. R., Quevedo, J., & Zhang, X. Y. (2020). Microglial Activation and Psychotic Disorders: Evidence from Pre-clinical and Clinical Studies. Current Topics in Behavioral Neurosciences, 44, 161–205. https://doi.org/10.1007/7854_2018_81

Bhaskar, S., Sinha, A., Banach, M., Mittoo, S., Weissert, R., Kass, J. S., Rajagopal, S., Pai, R., & Kutty, S. (2020). Cytokine Storm in COVID-19-Immunopathological Mechanisms, Clinical Considerations, and Therapeutic Approaches: The REPROGRAM Consortium Position Paper. Frontiers in Immunology, 11, 1648. https://doi.org/10.3389/fimmu.2020.01648

Chen, G., Wu, D., Guo, W., Cao, Y., Huang, D., Wang, H., Wang, T., Zhang, X., Chen, H., Yu, H., Zhang, X., Zhang, M., Wu, S., Song, J., Chen, T., Han, M., Li, S., Luo, X., Zhao, J., & Ning, Q. (2020). Clinical and immunological features of severe and moderate coronavirus disease 2019. The Journal of Clinical Investigation, 130(5), 2620–2629. https://doi.org/10.1172/JCI137244

Crespo-Facorro, B., Ortiz-Garcia de la Foz, V., Suarez-Pinilla, P., Valdizan, E. M., Perez-Iglesias, R., Amado-Senaris, J. A., Teresa Garcia-Unzueta, M., Labad, J., Correll, C., & Ayesa-Arriola, R. (2017). Effects of aripiprazole, quetiapine and ziprasidone on plasma prolactin levels in individuals with first episode nonaffective psychosis: Analysis of a randomized open-label 1year study. Schizophrenia Research, 189, 134–141. https://doi.org/10.1016/j.schres.2017.01.046

de Lusignan, S., Dorward, J., Correa, A., Jones, N., Akinyemi, O., Amirthalingam, G., Andrews, N., Byford, R., Dabrera, G., Elliot, A., Ellis, J., Ferreira, F., Lopez Bernal, J., Okusi, C., Ramsay, M., Sherlock, J., Smith, G., Williams, J., Howsam, G., … Hobbs, F. D. R. (2020). Risk factors for SARS-CoV-2 among patients in the Oxford Royal College of General Practitioners Research and Surveillance Centre primary care network: A cross-sectional study. The Lancet. Infectious Diseases, 20(9), 1034– 1042. https://doi.org/10.1016/S1473-3099(20)30371-6

Dinesh, A. A., Islam, J., Khan, J., Turkheimer, F., & Vernon, A. C. (2020). Effects of Antipsychotic Drugs: Cross Talk Between the Nervous and Innate Immune System. CNS Drugs. https://doi.org/10.1007/s40263-020-00765-x

Dobin, A., Davis, C. A., Schlesinger, F., Drenkow, J., Zaleski, C., Jha, S., Batut, P., Chaisson, M., & Gingeras, T. R. (2013). STAR: ultrafast universal RNA-seq aligner. Bioinformatics (Oxford, England), 29(1), 15–21. https://doi.org/10.1093/bioinformatics/bts635

Dyall, J., Gross, R., Kindrachuk, J., Johnson, R. F., Olinger, G. G. J., Hensley, L. E., Frieman, M. B., & Jahrling, P. B. (2017). Middle East Respiratory Syndrome and Severe Acute Respiratory Syndrome: Current Therapeutic Options and Potential Targets for Novel Therapies. Drugs, 77(18), 1935–1966. https://doi.org/10.1007/s40265-017-0830-1

Elhusseiny, K. M., Abd-Elhay, F. A.-E., & Kamel, M. G. (2020). Possible therapeutic agents for COVID-19: A comprehensive review. Expert Review of Anti-Infective Therapy, 18(10), 1005–1020. https://doi.org/10.1080/14787210.2020.1782742

Geer, L. Y., Marchler-Bauer, A., Geer, R. C., Han, L., He, J., He, S., Liu, C., Shi, W., & Bryant, S. H. (2010). The NCBI BioSystems database. Nucleic Acids Research, 38(Database issue), D492–496. https://doi.org/10.1093/nar/gkp858

Giridharan, V. V., Scaini, G., Colpo, G. D., Doifode, T., Pinjari, O. F., Teixeira, A. L., Petronilho, F., Macêdo, D., Quevedo, J., & Barichello, T. (2020). Clozapine Prevents Poly (I:C) Induced Inflammation by Modulating NLRP3 Pathway in Microglial Cells. Cells, 9(3). https://doi.org/10.3390/cells9030577

Guy, R. K., DiPaola, R. S., Romanelli, F., & Dutch, R. E. (2020). Rapid repurposing of drugs for COVID-19. Science (New York, N.Y.), 368(6493), 829–830. https://doi.org/10.1126/science.abb9332

Harrow, J., Frankish, A., Gonzalez, J. M., Tapanari, E., Diekhans, M., Kokocinski, F., Aken, B. L., Barrell, D., Zadissa, A., Searle, S., Barnes, I., Bignell, A., Boychenko, V., Hunt, T., Kay, M., Mukherjee, G., Rajan, J., Despacio-Reyes, G., Saunders, G., … Hubbard, T. J. (2012). GENCODE: the reference human genome annotation for The ENCODE Project. Genome Research, 22(9), 1760–1774. https://doi.org/10.1101/gr.135350.111

Juncal-Ruiz, M., Riesco-Davila, L., Ortiz-Garcia de la Foz, V., Martinez-Garcia, O., Ramirez-Bonilla, M., Ocejo-Vinals, J. G., Leza, J. C., Lopez-Hoyos, M., & Crespo-Facorro, B. (2018). Comparison of the anti-inflammatory effect of aripiprazole and risperidone in drug-naive first episode psychosis individuals: A 3months randomized study. Schizophrenia Research, 202, 226–233. https://doi.org/10.1016/j.schres.2018.06.039

Kato, T., Monji, A., Hashioka, S., & Kanba, S. (2007). Risperidone significantly inhibits interferon-gamma-induced microglial activation in vitro. Schizophrenia Research, 92(1–3), 108–115. https://doi.org/10.1016/j.schres.2007.01.019

Law, C. W., Chen, Y., Shi, W., & Smyth, G. K. (2014). voom: Precision weights unlock linear model analysis tools for RNA-seq read counts. Genome Biology, 15(2), R29. https://doi.org/10.1186/gb-2014-15-2-r29

Love, M. I., Huber, W., & Anders, S. (2014). Moderated estimation of fold change and dispersion for RNA-seq data with DESeq2. Genome Biology, 15(12), 550. https://doi.org/10.1186/s13059-014-0550-8

Luo, W., & Brouwer, C. (2013). Pathview: An R/Bioconductor package for pathway-based data integration and visualization. Bioinformatics (Oxford, England), 29(14), 1830– 1831. https://doi.org/10.1093/bioinformatics/btt285

Mangalmurti, N., & Hunter, C. A. (2020). Cytokine Storms: Understanding COVID-19. Immunity, 53(1), 19–25. https://doi.org/10.1016/j.immuni.2020.06.017

Manjili, R. H., Zarei, M., Habibi, M., & Manjili, M. H. (2020). COVID-19 as an Acute Inflammatory Disease. Journal of Immunology (Baltimore, Md.L: 1950), 205(1), 12– 19. https://doi.org/10.4049/jimmunol.2000413

Mayoral Van-Son et al. (2020). Comparison of aripiprazole and risperidone effectiveness in first episode non-affective psychosis: Rationale and design of a prospective, randomized, 3-phase, investigator-initiated study (PAFIP-3). Revista De Psiquiatría y salud mental. Accepted. November 2020

Obuchowicz, E., Bielecka-Wajdman, A. M., Paul-Samojedny, M., & Nowacka, M. (2017). Different influence of antipsychotics on the balance between pro- and anti-inflammatory cytokines depends on glia activation: An in vitro study. Cytokine, 94, 37–44. https://doi.org/10.1016/j.cyto.2017.04.004

Prieto, C., & Barrios, D. (2019). RaNA-Seq: Interactive RNA-Seq analysis from FASTQ files to functional analysis. Bioinformatics (Oxford, England). https://doi.org/10.1093/bioinformatics/btz854

Radhakrishnan, R., Kaser, M., & Guloksuz, S. (2017). The Link Between the Immune System, Environment, and Psychosis. Schizophrenia Bulletin, 43(4), 693–697. https://doi.org/10.1093/schbul/sbx057

Riva, G., Nasillo, V., Tagliafico, E., Trenti, T., Comoli, P., & Luppi, M. (2020). COVID-19: More than a cytokine storm. Critical Care (London, England), 24(1), 549. https://doi.org/10.1186/s13054-020-03267-w

Riva, L., Yuan, S., Yin, X., Martin-Sancho, L., Matsunaga, N., Pache, L., Burgstaller-Muehlbacher, S., De Jesus, P. D., Teriete, P., Hull, M. V., Chang, M. W., Chan, J. F.-W., Cao, J., Poon, V. K.-M., Herbert, K. M., Cheng, K., Nguyen, T.-T. H., Rubanov, A., Pu, Y., … Chanda, S. K. (2020). Discovery of SARS-CoV-2 antiviral drugs through large-scale compound repurposing. Nature, 586(7827), 113–119. https://doi.org/10.1038/s41586-020-2577-1

Rubino, S., Kelvin, N., Bermejo-Martin, J. F., & Kelvin, D. (2020). As COVID-19 cases, deaths and fatality rates surge in Italy, underlying causes require investigation. Journal of Infection in Developing Countries, 14(3), 265–267. https://doi.org/10.3855/jidc.12734

Smyth, G. K., Michaud, J., & Scott, H. S. (2005). Use of within-array replicate spots for assessing differential expression in microarray experiments. Bioinformatics (Oxford, England), 21(9), 2067–2075. https://doi.org/10.1093/bioinformatics/bti270

Vabret, N., Britton, G. J., Gruber, C., Hegde, S., Kim, J., Kuksin, M., Levantovsky, R., Malle, L., Moreira, A., Park, M. D., Pia, L., Risson, E., Saffern, M., Salomé, B., Esai Selvan, M., Spindler, M. P., Tan, J., van der Heide, V., Gregory, J. K., … Samstein, R. M. (2020). Immunology of COVID-19: Current State of the Science. Immunity, 52(6), 910–941. https://doi.org/10.1016/j.immuni.2020.05.002

Van den Berg, D. F., & Te Velde, A. A. (2020). Severe COVID-19: NLRP3 Inflammasome Dysregulated. Frontiers in Immunology, 11, 1580. https://doi.org/10.3389/fimmu.2020.01580

Van der Meer, D., Pinzón-Espinosa, J., Lin, B. D., Tijdink, J. K., Vinkers, C. H., Guloksuz, S., & Luykx, J. J. (2020). Associations between psychiatric disorders, COVID-19 testing probability and COVID-19 testing results: Findings from a population-based study. BJPsych Open, 6(5), e87. https://doi.org/10.1192/bjo.2020.75

Wang, Y., Song, F., Zhu, J., Zhang, S., Yang, Y., Chen, T., Tang, B., Dong, L., Ding, N., Zhang, Q., Bai, Z., Dong, X., Chen, H., Sun, M., Zhai, S., Sun, Y., Yu, L., Lan, L., Xiao, J., … Zhao, W. (2017). GSA: Genome Sequence Archive<sup/>. Genomics, Proteomics & Bioinformatics, 15(1), 14–18. https://doi.org/10.1016/j.gpb.2017.01.001

Weston, S., Coleman, C. M., Haupt, R., Logue, J., Matthews, K., Li, Y., Reyes, H. M., Weiss, S. R., & Frieman, M. B. (2020). Broad Anti-coronavirus Activity of Food and Drug Administration-Approved Drugs against SARS-CoV-2 In Vitro and SARS-CoV In Vivo. Journal of Virology, 94(21). https://doi.org/10.1128/JVI.01218-20

Wu, C., Chen, X., Cai, Y., Xia, J., Zhou, X., Xu, S., Huang, H., Zhang, L., Zhou, X., Du, C., Zhang, Y., Song, J., Wang, S., Chao, Y., Yang, Z., Xu, J., Zhou, X., Chen, D., Xiong, W., … Song, Y. (2020). Risk Factors Associated With Acute Respiratory Distress Syndrome and Death in Patients With Coronavirus Disease 2019 Pneumonia in Wuhan, China. JAMA Internal Medicine, 180(7), 934–943. https://doi.org/10.1001/jamainternmed.2020.0994

Xiong, Y., Liu, Y., Cao, L., Wang, D., Guo, M., Jiang, A., Guo, D., Hu, W., Yang, J., Tang, Z., Wu, H., Lin, Y., Zhang, M., Zhang, Q., Shi, M., Liu, Y., Zhou, Y., Lan, K., & Chen, Y. (2020). Transcriptomic characteristics of bronchoalveolar lavage fluid and peripheral blood mononuclear cells in COVID-19 patients. Emerging Microbes & Infections, 9(1), 761–770. https://doi.org/10.1080/22221751.2020.1747363

Zeng, Z., Yu, H., Chen, H., Qi, W., Chen, L., Chen, G., Yan, W., Chen, T., Ning, Q., Han, M., & Wu, D. (2020). Longitudinal changes of inflammatory parameters and their correlation with disease severity and outcomes in patients with COVID-19 from Wuhan, China. Critical Care (London, England), 24(1), 525. https://doi.org/10.1186/s13054-020-03255-0

Zhou, F., Yu, T., Du, R., Fan, G., Liu, Y., Liu, Z., Xiang, J., Wang, Y., Song, B., Gu, X., Guan, L., Wei, Y., Li, H., Wu, X., Xu, J., Tu, S., Zhang, Y., Chen, H., & Cao, B. (2020). Clinical course and risk factors for mortality of adult inpatients with COVID-19 in Wuhan, China: A retrospective cohort study. Lancet (London, England), 395(10229), 1054–1062. https://doi.org/10.1016/S0140-6736(20)30566-3

